# Topological data analysis model for the spread of the coronavirus

**DOI:** 10.1101/2020.08.13.20174326

**Authors:** Yiran Chen, Ismar Volić

**Author notes:** URL: ivolic.wellesley.edu.

## Abstract

We apply topological data analysis, specifically the Mapper algorithm, to the U.S. COVID-19 data. The resulting Mapper graphs provide visualizations of the pandemic that are more complete than those supplied by other, more standard methods. They encode a variety of geometric features of the data cloud created from geographic information, time progression, and the number of COVID-19 cases. They reflect the development of the pandemic across all of the U.S. and capture the growth rates as well as the regional prominence of hot-spots. The Mapper graphs allow for easy comparisons across time and space and have the potential of becoming a useful predictive tool for the spread of the coronavirus.

## 1. Introduction

Topological data analysis (TDA) is a method for understanding data clouds that attempts to gain insight into the data by treating it as a geometric object and extracting information based on its “shape”. There are several TDA instruments available, and the one we use in this paper is called the *Mapper algorithm*. Introduced by Singh, Mémoli, and Carlsson [14], Mapper is a way to simplify data while preserving many of its topological features. The idea is to project complicated data in a way that makes the projection tractable, then use topology to cover the projection with certain sets, and then look at the preimages of those sets under the projection map. This information is then used to construct a graph that retains many of the topological information about the original data set such as clustering, connectedness, 1-dimensional holes, etc. This graph lends itself more readily to analysis than the original data set, but is still complicated enough to provide more insight than some other dimension-reduction and clustering techniques such as principal component analysis. Mapper has been used in a variety of situations such as breast cancer data [10], image processing [13], and patent development [7].^1^ For overviews on Mapper and TDA more generally, see [1, 2].

In this paper, we apply the Mapper algorithm to the United States COVID-19 data. We consider the 4-dimensional data cloud consisting of the longitudes and latitudes of the U.S. counties and territories, number of days elapsed in the pandemic (starting on January 22, 2020), and the number of cumulative COVID-19 cases recorded by that day in each county.

The resulting Mapper graphs are rich in information and more complete than other more standard methods of visualization. They reflect important aspects about the development and spread of the COVID-19 pandemic across the U.S. and capture dramatic growth rates, trends, and regional prominence of hot-spots. In particular, compared with traditional ways of data visualization that similarly provide real-time monitoring of the spread of COVID-19 across the U.S. (e.g. Johns Hopkins University’s COVID-19 dashboard), Mapper visualization has the following advantages:

- It captures the entire developmental process of the outbreak in the U.S., including the emergence of hot-spots across time. Instead of displaying a plain snapshot of all the COVID-19 cases in the U.S. at certain point in time or a time series data for a single location, a Mapper representation captures the evolution of the spread in the entire country.
- The Mapper graph makes it easy to compare data across time and space. A certain U.S. county’s position in the graph is in part determined relative to the surrounding counties so, when irregular patterns appear in the visualization, it is because the data it represents has some volatility in comparison with its neighboring counties. This makes it easier to spot regional hot-spots while retaining an overarching view of the United States.

In Section 2.1, we give background on the Mapper algorithm, trying to keep the technical aspects to the minimum for clarity and readability. Sections 2.2 and 2.3 explain how we apply the Mapper algorithm to the U.S. COVID-19 data.

Section 3 contains the analysis of the various Mapper images we obtain. We discuss the meaning of connected components in Section 3.1 and explain how each of the coordinates - time, geography, and the number of cases - influence the connectedness of the graph. We turn to the other most prominent feature of the Mapper, its branches, in Section 3.2, and discuss how these structures indicate the appearance and development of COVID-19 hot-spots. Finally in Section 3.3, we take a look at the evolution of the Mapper with respect to time and examine how the succession of graphs provides useful information about the development of the pandemic across time and space.

In Section 4, we make some remarks about the many ways in which the work in this paper could be extended and generalized. For example, the Mapper could be overlaid with dates of stay-at-home orders so that the effectiveness and timeliness of such directives could be assessed. Socio-economic factors could also be read into the Mapper so that the COVID-19 spread could be correlated with such data. In addition, our analysis can be replicated for any region in the world, and, for some of them, travel restrictions between countries and border closings could also be incorporated into the data to gauge their efficacy. In addition, the branches that indicate hot-spots could be given more rigorous graph-theoretic structure that could lead to a robust predictive model. Finally, another TDA tool called *persistent homology* could be employed and used in conjunction with Mapper to gain an even deeper understanding of the spread of the COVID-19 pandemic.

Due to its relatively recent emergence, topological data analysis has not yet been applied extensively to epidemiology. The idea that this methodology could be useful in the study of viral evolution was put forth in [3]. The paper [15] provides a general framework for using TDA to study contagion across networks. Applications to particular diseases can be found in [4], which studies the spread of influenza, and [8], which does the same for Zika. All of the above papers use the persistent homology arm of TDA. The Mapper algorithm does not appear to have been used in the context of epidemiology yet, except in [5], which uses the *Ball Mapper* to examine economic and COVID-19 case data in England; this also seems to be the only article that studies the coronavirus pandemic through TDA to date.

### 1.1. Acknowledgements

The second author would like to thank the Simons Foundation for its sup-port.

## 2. Methods

### 2.1. The Mapper algorithm

Given a potentially high-dimensional data set *X* (each data point is possibly a vector in some high-dimensional Euclidean space), Mapper replaces it by a graph consisting of nodes and edges that retain information about some of the original geometric features of the data (more generally, Mapper creates a higher-dimensional version of a graph called a *simplicial complex*, but we will stick to graphs in this paper). Not only are various important features such as components and holes preserved, but Mapper’s visual representation of the data can often reveal new structures and insights like flares or branches that other, more traditional statistical methods, cannot.

To implement Mapper:

1. Use some projection function (also called *filter* or *lens*) *f: X →* ℝ*^d^* to map the dataset into some Euclidean space ℝ*^d^*.
2. Cover *f*(*X*), the image of the dataset in ℝ^d^, by a collection of overlapping open sets

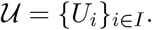 Here *I* is some finite indexing set. Sets *U_i_* are also sometimes called *bins*.
3. Apply some clustering algorithm to each preimage *f*^−1^(*U_i_*) (preimages are also called *fibers*). This produces *i_j_* clusters 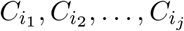 in the ith preimage.
4. Create a graph whose vertices or nodes consist of the set of all clusters

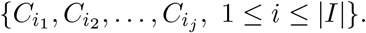 An (unoriented) edge between nodes 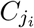 and 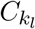 is added if and only if

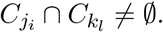

This collection of nodes and edges is the Mapper graph, denoted by *M*(*X*, 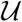, *f*).^2^

Figure 1 illustrates this procedure on a simple dataset *X* that lives in ℝ^2^, with the projection function *f*: *X* → ℝ mapping to the real line by forgetting the first coordinate.

**Figure 1.**
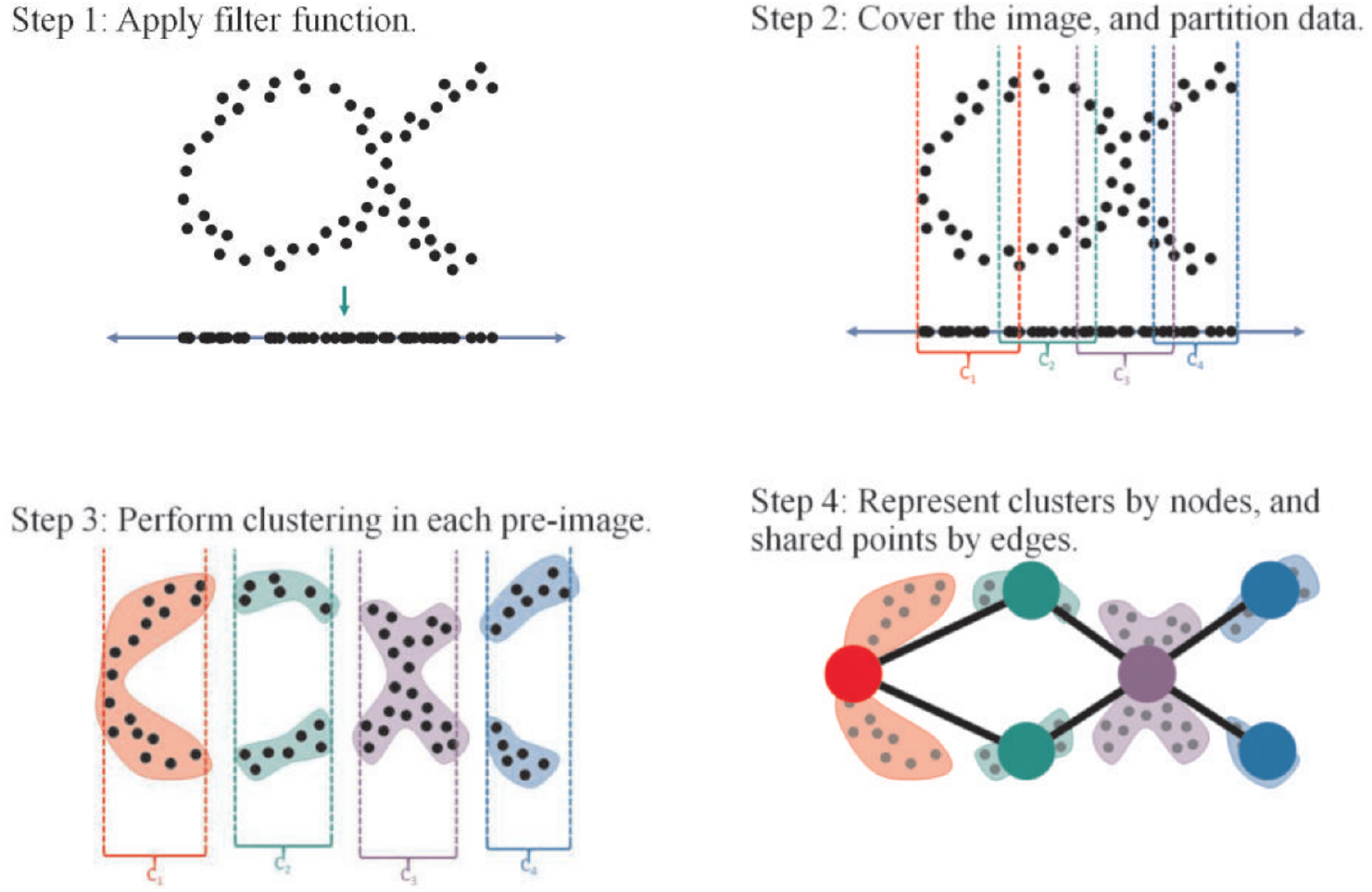
An illustration of the Mapper algorithm. Image Source: Escolar, Hiraoka, Igami, Ozcan [7].

The projection function depends on the context and on the aspect of the data that is deemed most relevant. It could simply forget coordinates, but most useful projections are statistically more meaningful, such as the kernel density estimator, distance to measure, or graph Laplacian. In the applications of Mapper to the study of cancer, for example, a Healthy State Model which decomposes tumor cells into the “disease” and “healthy” components is used to project to the disease part [10]. The domain of the projection is often ℝ, i.e. the projection simply assigns a real number to each data vector.

The image *f*(*X*) is usually covered by hypercubes (products of intervals), but open covers of any shape are a priori allowed. The number and the amount of overlap of the open sets determines many features of the graph, including the number of its connected components.

The task of a clustering algorithm is to determine which subsets of data points in each preimage appear to be closer to each other than to others. This is essentially the discrete version of identifying the connected components of a topological space. Clustering can be done by selecting an appropriate metric or distance function (Euclidean, correlation, etc.) on the data and using it to decide the “closeness” via a procedure such as *single-linkage hierarchical clustering*. In fact, there is no need for the distance function to satisfy the triangle inequality, so technically only a *semimetric* is required. Clustering can also be done in the image *f*(*X*), but some information loss due to projection is possible.

The size of the nodes can also be regulated and it corresponds to the number of data points in that cluster. Nodes can also be colored according to some chosen characteristics of the data. For example, in the applications of Mapper to breast cancer data [10], the nodes are colored according to survival rates.

Note that there are many choices that are left to the user - the projection function, the number of open sets in the cover of *f*(*X*) and the amount of their overlap, and the clustering algorithm. This ad hoc nature of the algorithm is in many ways its advantage because it allows for great generality and flexibility in the types of data that can be treated and the questions that can be asked about it.

Mapper is an unsupervised data analysis method, but is more subtle than other ones such as principal component analysis. It performs dimensionality reduction and clustering while preserving the shape of data. This is possible because Mapper clusters locally but then extrapolates back to global data via the edges. The graph is often a lot easier to interpret than other visualizations like scatterplots and it effectively captures many facets of the original, high-dimensional data.

There are various free technical implementations of the Mapper algorithm, such as Python Mapper [9], TDAmapper (ℝ package) [12], and Kepler Mapper (which we use in this paper) [16].

### 2.2. COVID-19 data

The data we use in this study comes from the COVID-19 Data Repository by the Center for Systems Science and Engineering (CSSE) at Johns Hopkins University.^3^ Specifically, we use a collection of time series data on the number of confirmed COVID-19 cases reported by 3155 U.S. counties (and territories) starting on 1/22/20. The end date for most of our analysis will be 6/19/20, but some of it will go up to 7/24/20 (see Figures 9 and 10 in particular). Each data point encodes information on the geographic location of a certain U.S. county, a date, and the number of cumulative confirmed COVID-19 cases reported in that county on that date.

More precisely, suppose a U.S. county is located at latitude *x* and longitude *y*. Here *x* is positive when it represents a northern latitude and negative when it represents a southern one (e.g. American Samoa). Similarly *y* is positive if it is an eastern longitude (e.g. Guam) and negative otherwise.

Let the coordinate *w* encode the date information as relative to the first date in our time series, namely 1/22/20, which is itself given value 0. Lastly, let *z* be the number of cumulative confirmed COVID-19 cases in that county on date *w*. Then our cloud consists of data points

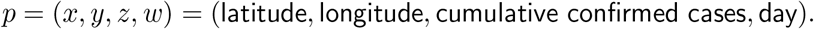

For example, King County in Washington state is located at roughly 47°*N*, 122°*W* and reported 7472 cumulative confirmed COVID-19 cases on 5/17/20. In our original point cloud, this data point is thus encoded as

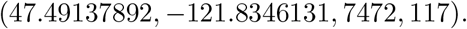

The entire data cloud up to 5/17/20 will then contain

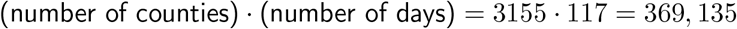

vectors of length 4, i.e. elements of ℝ^4^.

#### 2.2.1. Normalization

Since the four coordinates of our data vector use varying systems of measurement, which cause their numerical values to have different orders of magnitude, modeling them directly into a 4-dimensional Euclidean space to serve as the basis for Mapper clustering means that the four coordinates will take on disproportionate weights in determining the shape of the Mapper graph. More specifically, coordinates that have relatively small numerical values, such as the time coordinate, will barely be evident in the Mapper graph. We hence need to offset the distortion effect induced by the numerical discrepancies in the coordinates of our data point. We achieve this by separately scaling each coordinate of the input row vectors to column-wise unit norm. This means that if we square a chosen coordinate of all the scaled vectors and then sum them within its feature, the total would be 1.

From now on, we will be working with this normalized point cloud. Note that, as the time interval is modified (by changing the end date), the values of the coordinates of the vectors change under the normalization as well, i.e. the same vector might be normalized to different values after the time frame is extended to a later date.

### 2.3. Application of the Mapper algorithm to the COVID-19 data

Recall the general description of the Mapper algorithm from Section 2.1. To construct a Mapper graph for our point cloud X, we use the Python implementation of the Mapper, KeplerMapper by Van Veen and Saul [16], along with the following specifications:

1. The projection function is the identity map, namely *f*: *X* → ℝ^4^ simply sends a vector to itself. That is, since our data is not high-dimensional nor are we after any particular statistical features of the data cloud, there is no need to project it.
2. To cover the data (equivalently, the image of *f*), we use the standard Euclidean metric in ℝ^4^ and the default KeplerMapper cover procedure with Euclidean 4-dimensional cubes and the parameter *n* = 10. This means that the projections of the data cloud onto each of the axes in ℝ^4^ is covered by 10 overlapping intervals, and then each cube is formed as the cartesian product of those intervals. The degree of the overlap will be *δ* = 8%. These values are chosen because empirically they appear to give the most informative Mapper graphs. These parameters will be modified slightly for some of the pictures in Section 3.3.
3. For the clustering algorithm, we use the default DBSCAN^4^ clustering offered by KeplerMapper. The advantage of DBSCAN is that it identifies clusters of any shape (unlike other methods which only look for convex clusters).

The colors in our Mapper graphs do not have any mathematical meaning. The nodes are colored according to how the data is ordered, and this is done based on the geographic information, so that nearby counties are colored similarly. We can thus use the colors to add coherency among graphs and to help distinguish geographic locations.

A representative Mapper visualization of our data can be found in Figure 2. This is a visualization of the COVID-19 data from all 3155 U.S. counties and territories as of 6/19/20.

**Figure 2.**
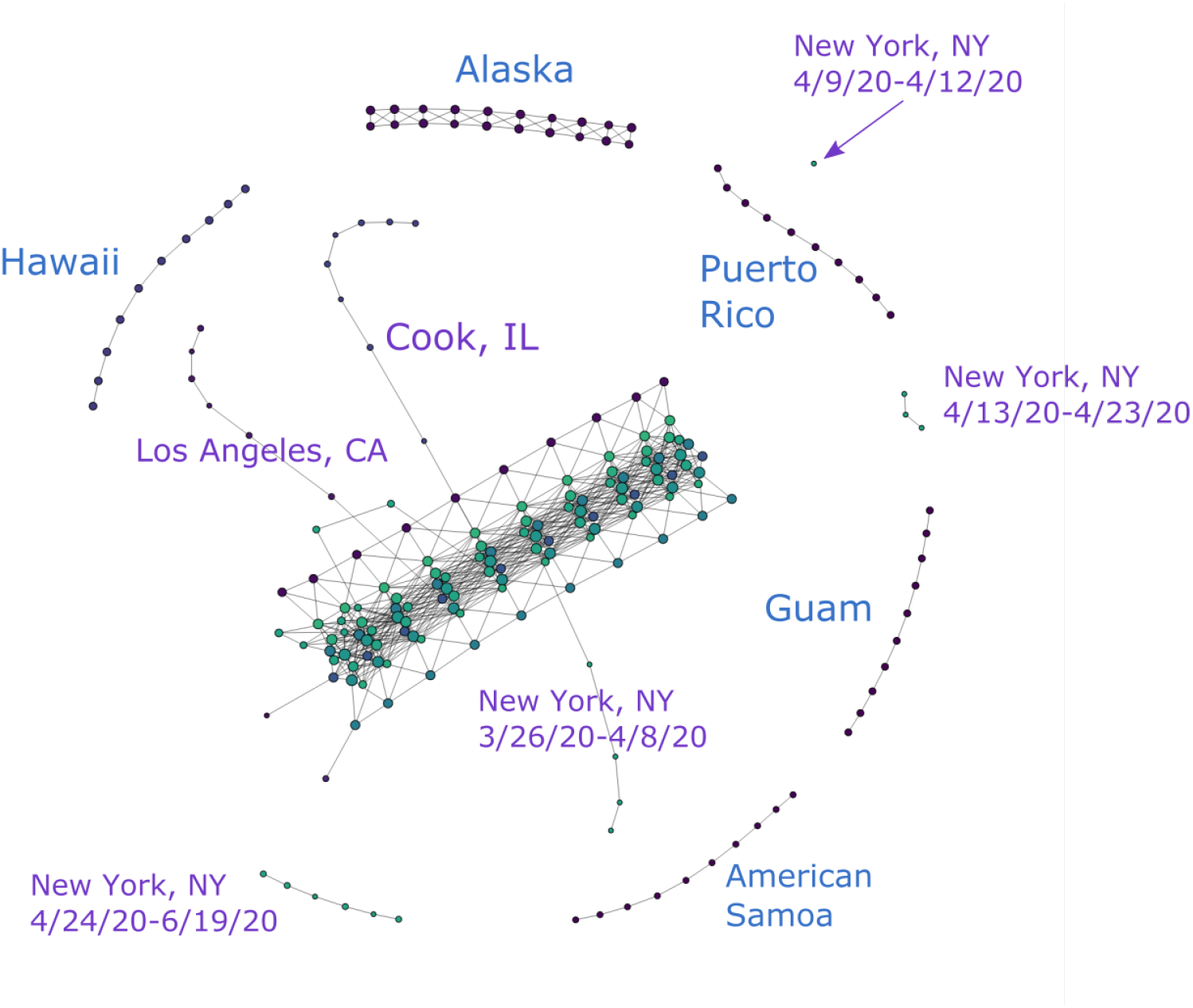
COVID-19 cases reported in the U.S. as of 6/19/20. (*n* = 10*, δ* = 8%)

Figures 3 and 4 show partial content of two representative nodes. The first node contains 25,824 data points. The geographic diversity of the counties in this node is evident from the number of bars of different colors in the top left. In contrast, the second node only has 31 data points, all of which come from New York State. Additionally, the size of each node is also indicative of the number of data points it contains.

**Figure 3.**
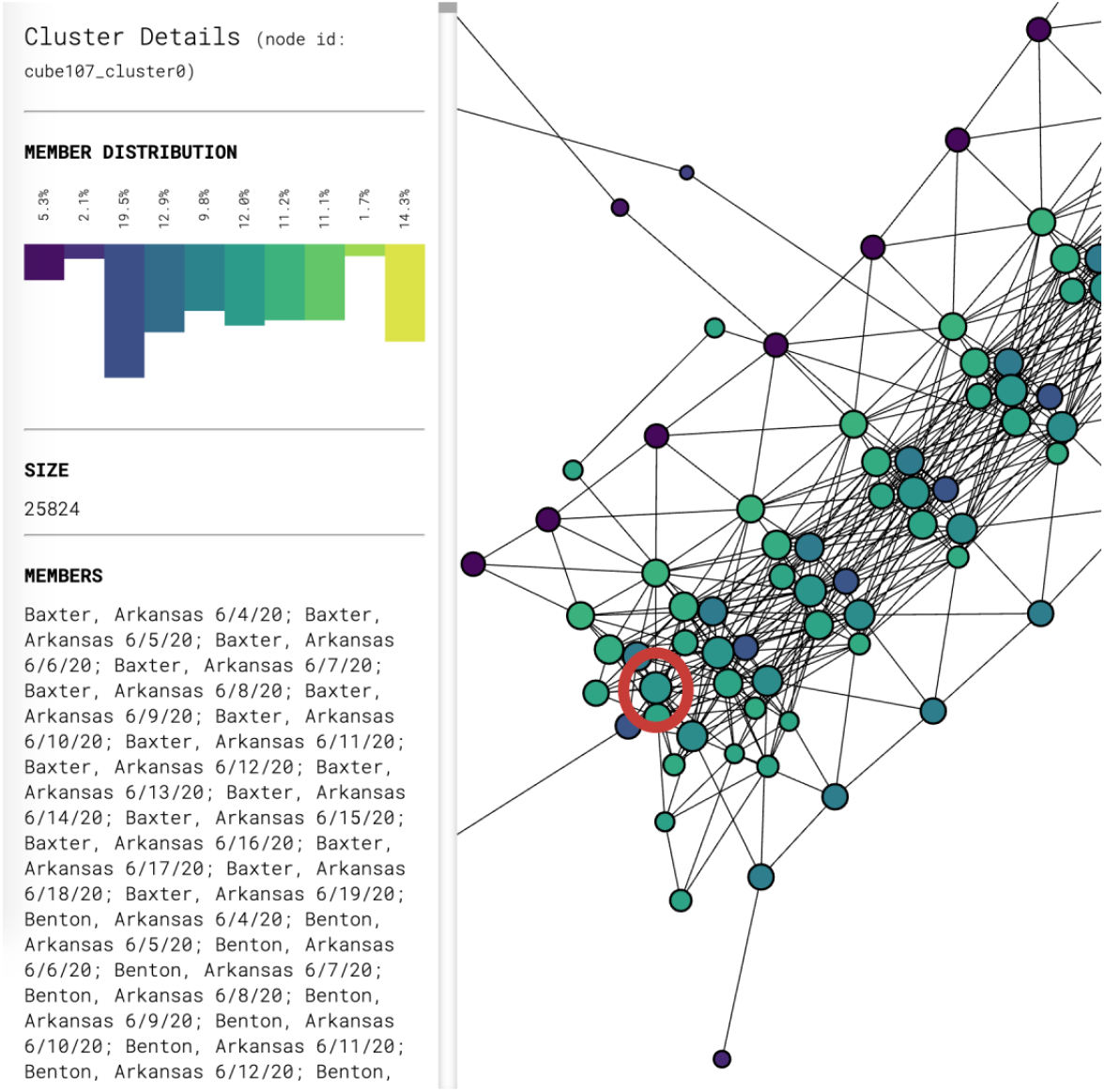
Breakdown of the data inside the node circled in red. Only the beginning of the list of the 25,824 data points in this node is shown.

**Figure 4.**
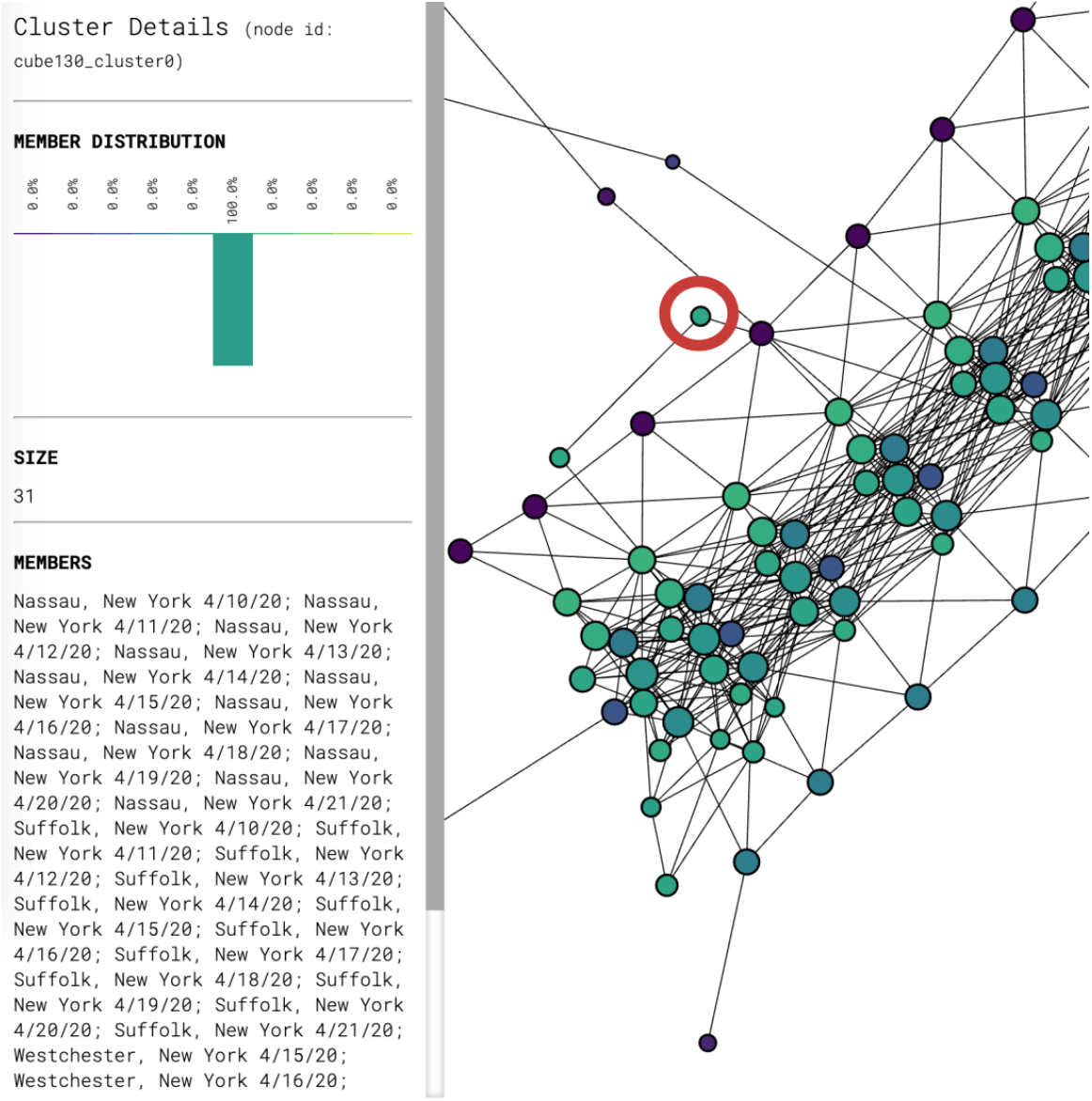
Breakdown of the data inside the node circled in red. The node contains 31 data points, all of which come from New York State.

#### 2.3.1. Filtering

As mentioned in Section 2.2.1, each of the four vectors consisting of a particular coordinate of each data point is scaled to unit norm. The resulting normalization for a data point is determined by the relative numerical significance of each coordinate within their own coordinate peers. This normalization is hence substantially affected by outliers in the data set, which can obscure a significant amount of data that would otherwise be numerically distinguishable. For example, looking at Figure 2, all counties in New York State, Los Angeles County in California and Cook County in Illinois have disproportionately large numbers of cumulative COVID-19 cases. They significantly increase the range of the data and are thus given more weight on the normalization process. As a result, their presence in the point cloud reduces the numerical significance of other data points and makes them rather indistinguishable in the Mapper graph.

To solve this issue, we filter our data set to produce different graphs either with or without these outliers. In the rest of the paper, we refer to data sets that include the aforementioned outlier counties as “unfiltered” and the ones without as “filtered”. For instance, Figure 2 makes it visually obvious that New York, Cook and Los Angeles “stand out” in the form of branches in comparison with rest of the data that congregate in the main trunk. On the other hand, Figure 5 provides the Mapper graph representing the same data set, but filtered to exclude data from Los Angeles, Cook, IL and the entire New York state. It is evident from this graph that more branches emerge as a result of filtering. This provides more helpful information on places that are not top-ranked in terms of total COVID-19 cases but still display worrisome trends or regional significance, such as King, WA and Maricopa, AZ.

**Figure 5.**
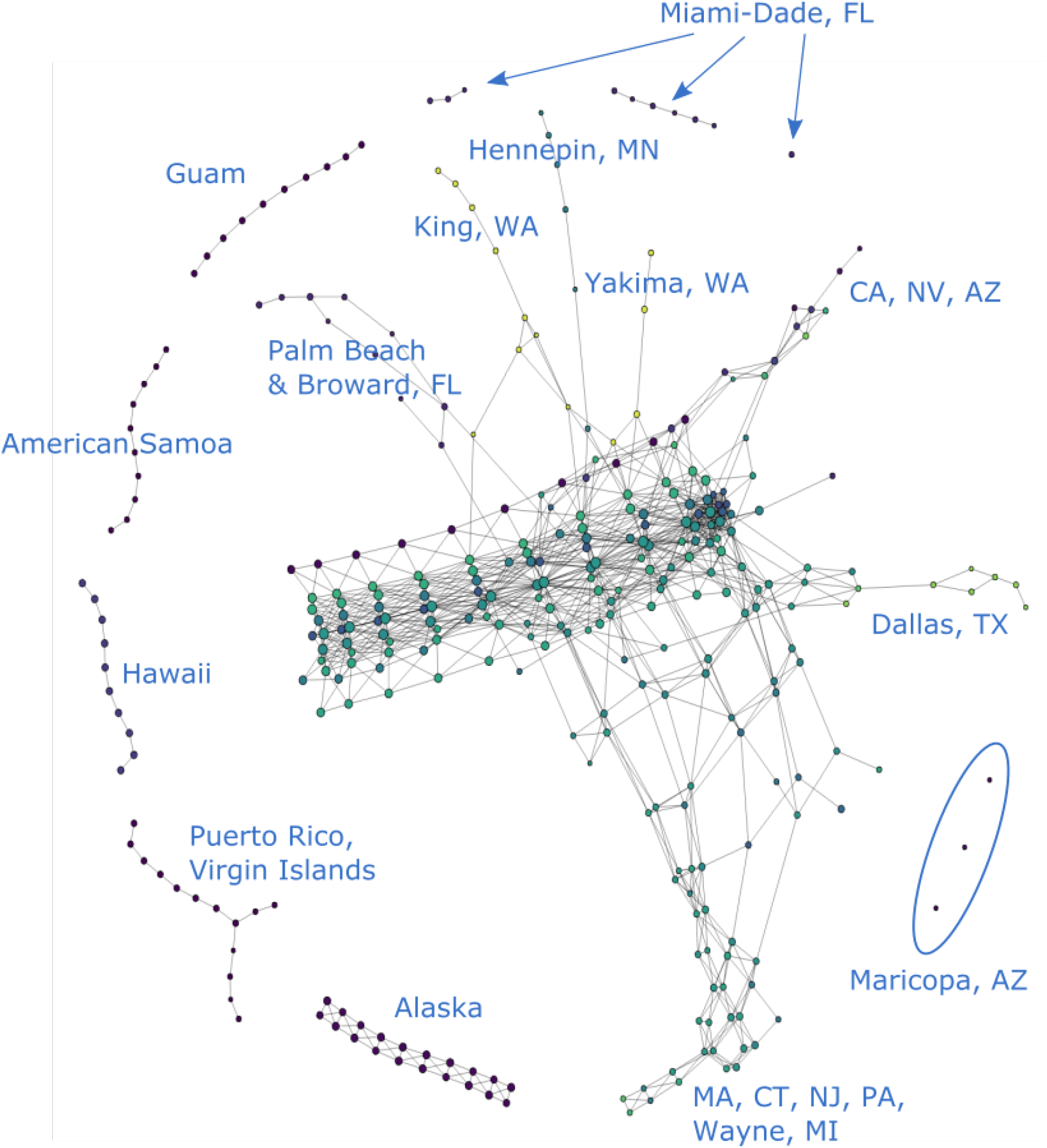
COVID-19 cases reported in the U.S. as of 6/19/20, excluding data from the entire New York State, Los Angeles County, CA and Cook County, IL. (*n* = 10*,δ* = 8%)

The rest of the paper works with both unfiltered and, for the most part, filtered data sets. We mostly use the former for New York-related analysis and the latter for other prominent U.S. COVID-19 clusters like the Maricopa County in Arizona.

For the filtered and unfiltered graphs that contain more recent dates (up to 7/24/20), see Figures 9 and 10.

## 3. Results

In this section, we introduce several significant topological features that could be observed from the Mapper graph, including components and branches, and illustrate what could be inferred from the presence and structure of these features.

### 3.1. Components

It is clear from Figure 2 that the original point cloud clusters into several disjoint components under the Mapper algorithm. Most of the data lives in the main “trunk,” but there are also several smaller isolated components. These disjoint components are formed either by a single isolated node or by a group of nodes connected by edges. Recall that an edge is created between two nodes when there is a data point living in both, and that a node is formed by covering the projections of the point cloud to each axis in ℝ^4^ with overlapping, equal-length intervals and then clustering the data points within each 4-dimensional cube determined by these intervals. Connected clusters of nodes are thus formed when the data in each node displays close similarity or proximity, and disjoint components emerge when they are numerically dissimilar, so that the data points are “far away,” i.e. there are gaps between the projections of data points into any of the four dimensions, resulting in empty overlaps between covers. Significant variations in any of the four coordinates among data points therefore break the nodes or clusters of nodes into disjoint components.

Because the projection of the point cloud is linear along the time dimension before the normalization, this distribution does not result in empty overlaps when covers are applied. Therefore, instead of breaking the Mapper into isolated components, the time dimension rather accounts in part for the connectedness features of the Mapper and, furthermore, to the internal distribution of nodes within each component. Therefore, we can usually explain the existence of isolated components by variations in the other three coordinates, which concern two factors:

a. Geographic variations of the counties represented by the data points; and
b. Variations in the number of cumulative confirmed cases reported by each county.

We will discuss the ramifications of these two factors below in Sections 3.1.2 and 3.1.3. Before that, we will say more about how the time component plays a role in keeping the nodes connected and in shaping the internal structure of most components.

#### 3.1.1. Time

The time coordinate is unique in the sense that its values follow a linear progression before normalization. Because of this linearity, normalized coordinates stay relatively dense and empty overlaps will thus almost never emerge when covers are applied to the projection of the data into the time axis, notwithstanding most choices of *n* and *δ*. This implies that time hardly explains the disconnectedness of the graph but in fact produces a considerable number of the edges between nodes and clusters.

Moreover, when covered by equal-length intervals, this dense distribution results in highly predictable, segmented internal structures within most clusters in the Mapper graph, so that the main trunk and most other substructures could even be seen as having been formed by weaving together clusters or nodes with data from successive time segments. To see this, let us first remove the time coordinate and produce a Mapper graph with data from a single day as in Figure 6. Without the time component, we can clearly observe the impact of geography on the shape of the graph. If we compare this graph with Figure 7, which is generated with the time coordinate included, we see that the latter graph could be regarded as having been formed by horizontally juxtaposing and connecting graphs produced on consecutive dates, each looking like Figure 6, and then applying the clustering algorithm along the time axis to further cluster the points.

**Figure 6.**
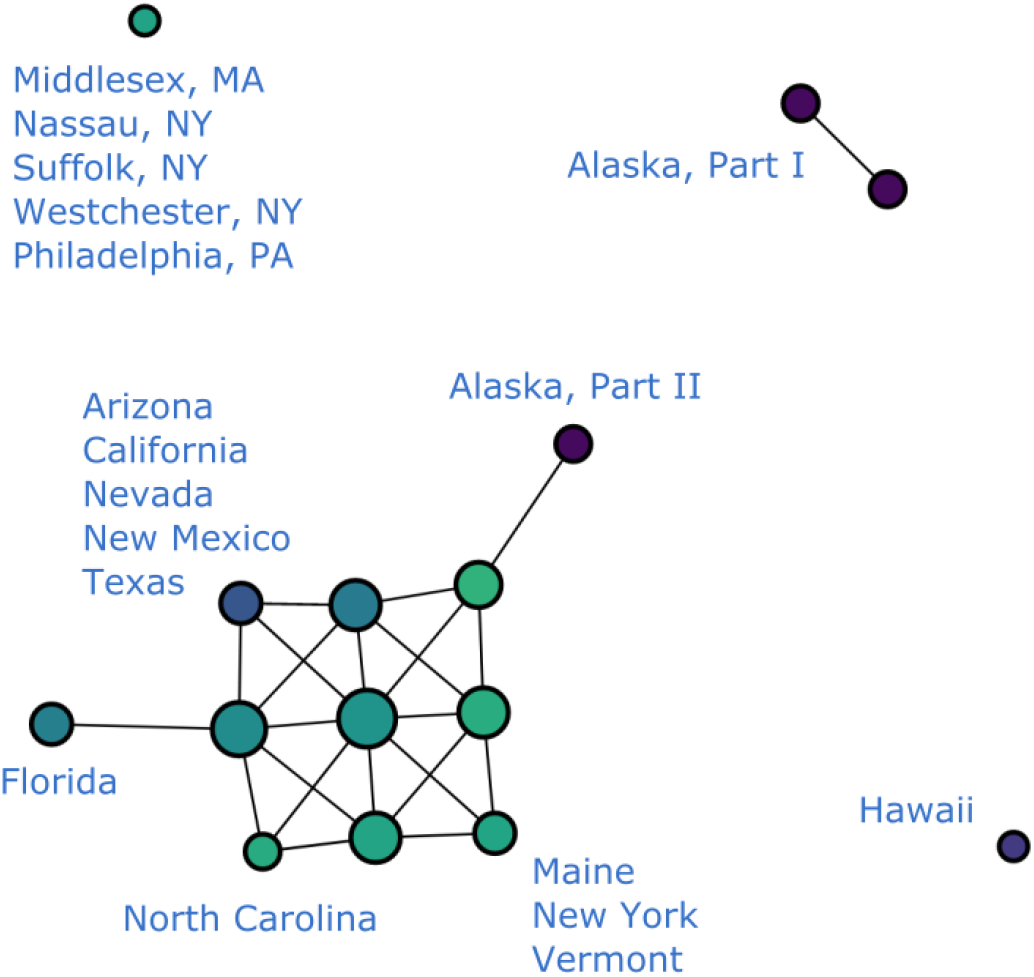
Cumulative COVID-19 cases reported in the U.S. on 7/1/20. (*n* = 10*,δ* = 8%) *Note*: Alaska, Part I houses the following Alaskan counties: Anchorage, Bethel, Denali, Fairbanks North Star, Kusilvak, Matanuska-Susitna, Nome, North Slope, Northwest Arctic, Southeast Fairbanks, Valdez-Cordova, Yukon-Koyukuk, Aleutians East, Bristol Bay, Dillingham, Kenai Peninsula, Kodiak Island, Lake and Peninsul, Yakutat, while Alaska, while Part II houses Aleutians West, Haines, Hoonah-Angoon, Juneau, Ketchikan Gateway, Petersburg, Prince of Wales-Hyder, Sitka, Skagway, and Wrangell.

**Figure 7.**
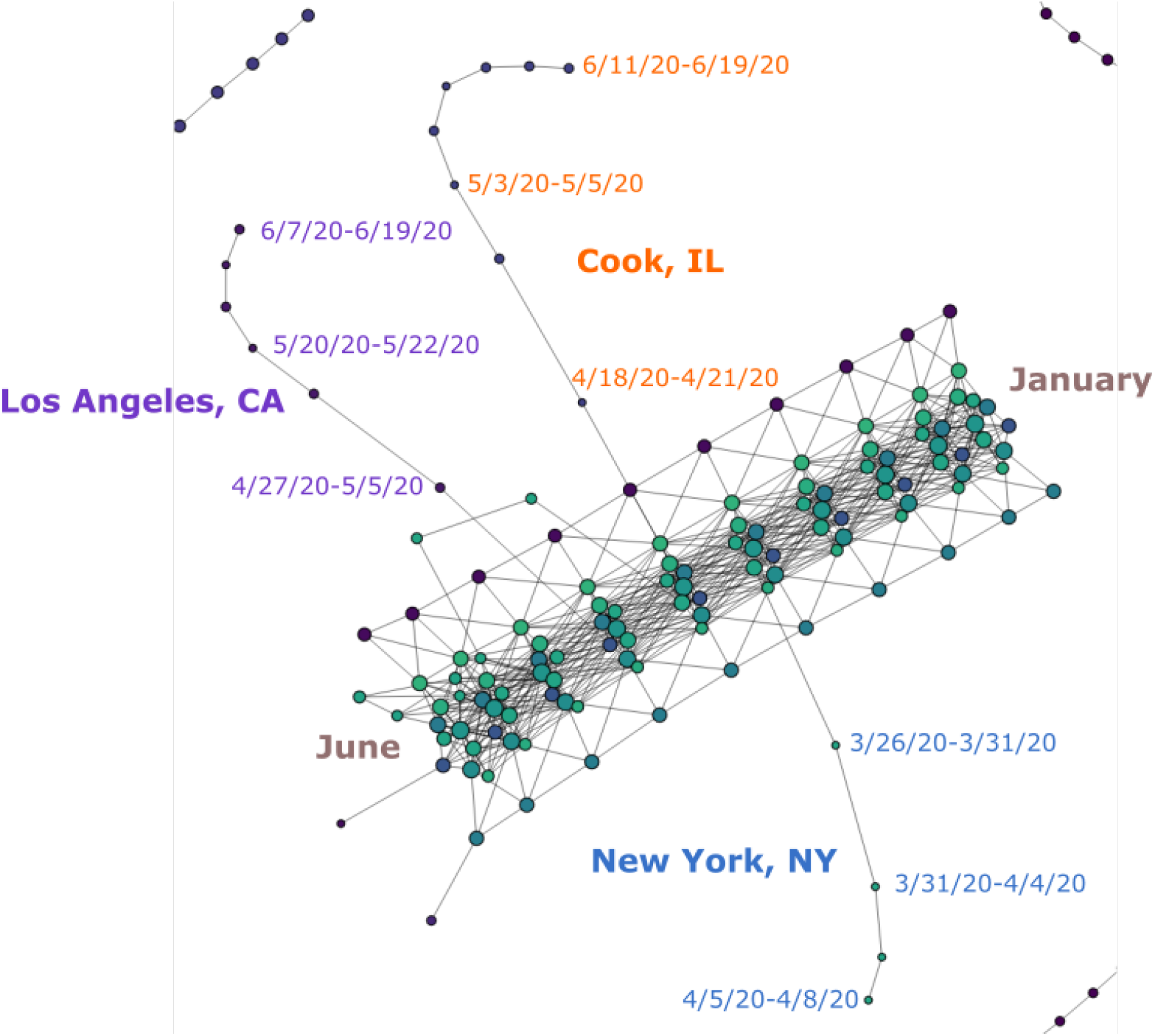
Main trunk and branches of Figure 2 showing time progression.

The result, as we can observe in Figure 7, is that our Mapper graph displays the evolution of cases through the progression of time in an intuitive manner, where the trunk and most other substructures appear to have “grown” in accordance with time data like the trunk of a tree, building up on data from the first few days in our data set. The right side of the trunk contains most of the U.S. counties at the beginning of the time period in January, and the time progresses toward the left. The left end of the trunk contains the data for June. The “branches” that stick out of the main trunk also follow this pattern. The nodes that make up the branches in Figure 7 are clearly arranged in accordance with the progression of time. Hence each branch represents the development of COVID-19 cases within a certain period of time. We will study these branches in depth in Section 3.2.

#### 3.1.2. Geography

Geographic information is an important factor that affects the distribution of the data throughout the Mapper graph and is readily reflected through this visualization. For example, the nodes representing Hawaii, Alaska, Guam, Puerto Rico and American Samoa in Figure 2 naturally form independent clusters. This is explained by their geographic separation from the U.S. mainland, which is for the most part represented in the Mapper graph as the main trunk. Specifically, the geographic separation for these regions is reflected in the data in the relative numerical disparity in the first two coordinates. With appropriate values of *n* and *δ*, this numerical disparity will give rise to disjoint clusters lying outside of the main trunk in the Mapper graph, as they do in Figure 2.

The geography is also more transparent when we remove the time coordinate and produce a Mapper graph for a single day only, as we do in Figure 6. The grouping of data points into clusters highly correlates with U.S. geography. However, we can also spot an isolated node in the graph representing several counties in New York, Pennsylvania and Massachusetts. This result should clearly not be attributed to geography and we thus know that geographic information is not deterministic to the shape of the Mapper graph. Since we have already removed the time component from our data before generating this graph, the only coordinate left that could explain this distribution is the number of cases reported.

#### 3.1.3. Number of COVID-19 cases

As explained in Section 2.2, the third coordinate of our data points records the cumulative number of COVID-19 cases reported daily from each county. In comparison with geographic and temporal information, this coordinate usually has the largest standard deviation among the four features.^5^ Before normalization, its numerical value can range from a few hundred for less-affected areas to several hundred thousand for hot-spots like the New York City, or, more recently, some areas in Florida and Texas. Therefore, this coordinate also adds the most variability into the shape of the Mapper graph.

This is mostly evident in two aspects. Similar to what happens with the geographic information, a large disparity in the numerical values of this coordinate can distribute data points into disjoint components outside of the main trunk. For example, we see in Figure 6 that there is an isolated node in the upper left corner whose presence could not be explained by geographic isolation. In fact, this node houses several counties with the highest numbers of cumulative COVID-19 cases reported by the time this graph was generated. The Mapper representations of such counties thus correspondingly stand out from those of the places that report only a mediocre number of cases. This type of distribution is also observed in ordinary Mapper graphs like Figure 2 where temporal data is included.

For the most part, however, the elevations of COVID-19 cases are reflected through the emergence of “branches” sticking out of the main trunk. Under this point of view, the free-floating clusters or nodes discussed above can be seen as fragmented segments of an otherwise connected branch. We will examine these branches in more detail next.

### 3.2. Branches

We see in Figure 2 that the data for Los Angeles, CA, Cook, IL and New York, NY presents itself in the form of chains of nodes branching off the main trunk. These three counties ranked top three in terms of COVID-19 cases reported by the time this graph was generated. As discussed in previous sections, because geographic and temporal data have relatively small standard deviation, they cause the graph to stay connected in these dimensions rather than producing features like these branches. This is evident in the “branch-free” part of the trunk, which represents data from early days of the outbreak when there is no significant disparity in the numbers of cases reported from different regions. It is therefore the high number of cases reported in these places that forces the nodes representing these counties to depart from the ones representing their geographic neighbors.

With appropriate choices of *n* and *δ*, we can thus produce Mapper graphs where regions with relatively large numbers of COVID-19 cases are no longer bound to others through edges in the geographic dimensions but remain connected solely in the time dimension, resulting in the branching feature that we see in Figure 2.^6^ Hence, the growth of these branches tracks the incremental increase of cases in these counties.

Additionally, because different segments of the main trunk are built with their own timestamps as discussed in Section 3.1.1, the place where each branch starts to grow is therefore indicative of the onset of the outbreak that it represents. That is to say, a branch that emerges later in the trunk signifies a more recent outbreak. In this way, we can see that the Mapper graphs encodes the development of the pandemic in these hot-spots in an intuitive manner. In the following sections, we will offer several case studies on various branches of different shape and elaborate on what could be learned from them.

#### 3.2.1. Segmented branches: New York and others

As we just explained, the emergence of branches signifies potential or existing COVID-19 hot-spots. However, some hot-spots like New York are more prominent than others; this empirical phenomenon also has a representation in the Mapper graph. Namely, looking at Figure 2, what we observe is that the branch representing data from the New York County is in fact broken into several segments, with each segment representing COVID-19 cases reported during a certain period of time. This distribution can be regarded as reflecting several stages in the development of the pandemic in New York during which several dramatic spikes of cases occurred.

In particular, 4/9/20, 4/13/20, and 4/24/20 are the critical dates underlying these disruptions in the branch. As is visible in Figure 8, these dates (labeled in red) correspond to either spikes in the number of daily new cases reported in New York County or a nadir before the next peak. Hence, the disruptions in the branch usually occur as a result of an extended period of elevated daily incidence, which forces the projections of the succeeding series of data points along the “number of cases” dimension to land in a hypercube far enough so that it breaks the continuity established by densely populated data points along the time dimension. Additionally, we notice that earlier peaks of daily new cases did not break up the branch. Instead, they contribute to the growth in the “length” of the branch by adding a new node. That is, several such peaks that occurred on 3/26/20, 3/31/20, and 4/4/20 (labeled in yellow) correspond to the critical dates that cause a new node to be added to the branch. Therefore, many noteworthy dates in the development of the pandemic in New York County are reflected in the graph either in the form of new nodes or disruptions of the branch. Several snapshots of the dynamic process through which segments of this branch gradually emerged can be found in Figure 13.

**Figure 8.**
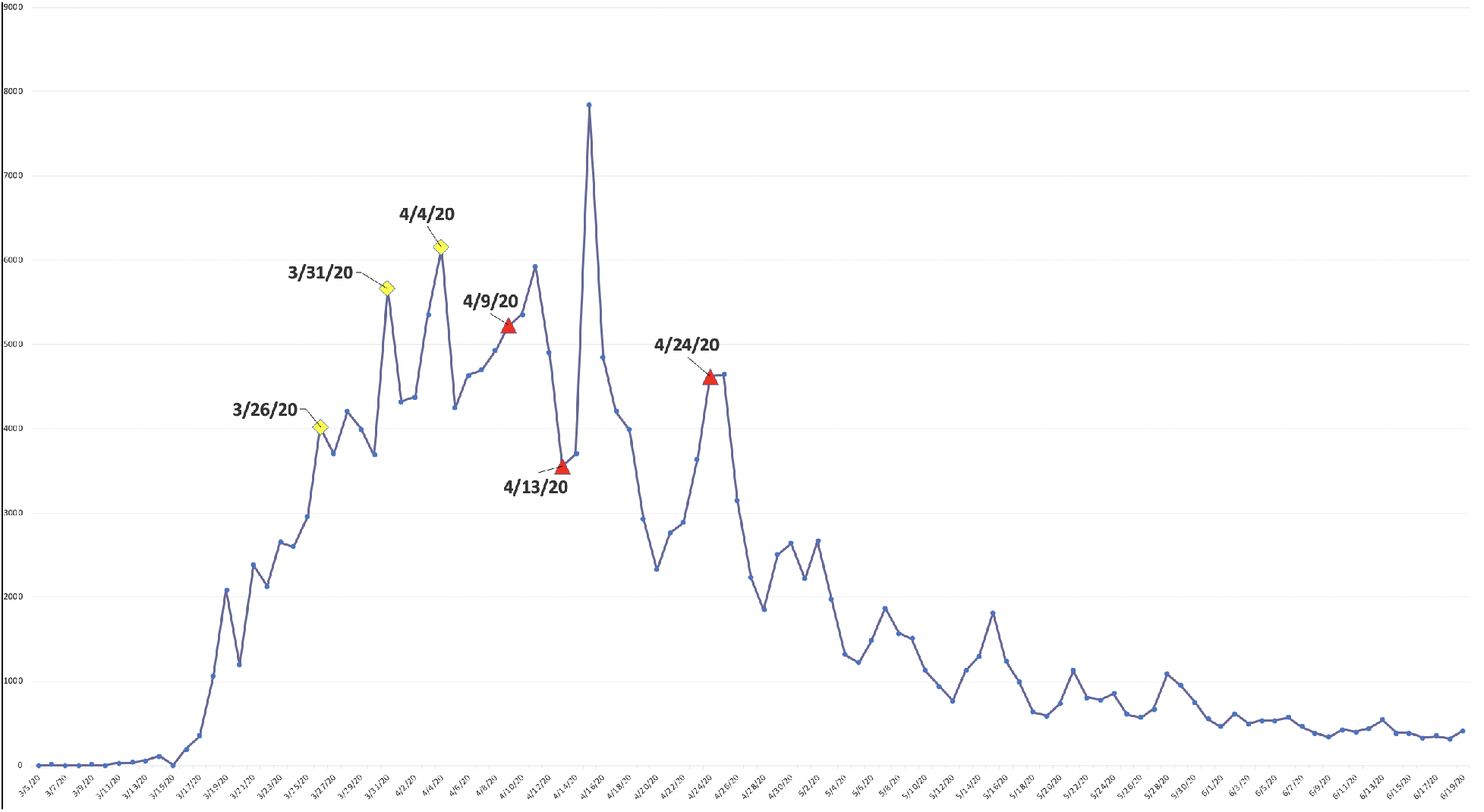
Daily new cases reported in New York, NY from 3/5/20 to 6/19/20.

Similarly, in the Mapper graphs generated from filtered data sets, some prominent COVID-19 hot-spots are also exposed in the form of segmented branches. They are Miami-Dade, FL and Maricopa, AZ, and can be seen in Figure 5. Similarly to New York, these places also experienced aggressive growths in daily incidence. Broken branches therefore generally symbolize high levels of daily growths and signify alarming trends in the regions they represent.

#### 3.2.2. Branch complex

A branch complex is formed when a number of closely-situated counties report similar high numbers of cases and display elevated daily incidence. In this case, each county is represented as a branch that departs from the main trunk. However, because of the geographic and temporal proximity of these outbreaks, nodes in their respective branches tend to be connected through the geographic dimension as well. This explains the various entangled branching structures emanating from the main trunk in Figure 5.^7^ For example, one can see the complex formed by a number of counties in the Northeast and Wayne, Michigan as well as a less complicated one formed by counties in California, Nevada, and Arizona. Another small complex is formed by two counties in Florida, i.e. Palm Beach and Broward.

Because of their prominence, these complexes tend to develop into trunks of their own so that a resurgence of the pandemic in these existing hot-spots will be reflected in the Mapper graph in the form of new branches that grow out of the branch complex instead of the main trunk. On the other hand, the emergence of a new hot-spot in nearby regions enlarges the complex by entangling a new branch into it.

#### 3.2.3. More recent hot-spots

As mentioned in Section 3.2, the timing of the onsets of regional flareups is indicated by the position of the resulting branches relative to the main trunk. For example, we can see from Figure 2 that the outbreak in New York occurred prior to the ones in Cook and Los Angeles, because its branch emerged earlier in time. We can similarly distinguish more recent clusters of outbreaks from older ones in this way. For instance, it is clear in Figure 5 that the outbreaks in Dallas, Texas and Nevada occurred after the ones in Palm Beach and Broward, Florida or in King, Washington as their branches sit closer to the end of the trunk that corresponds to later dates. They therefore represent a new generation of COVID-19 hot-spots, distinct from more mature ones in Northeast U.S. or Washington state.

To ensure the coherency in our analysis, we only studied graphs produced with data collected prior to 6/20/20 in previous sections. For a more recent update in these graphs, see Figures 9 and 10. It is noteworthy that these graphs make several new hot-spots readily visible. They include Maricopa County in Arizona, Harris and Dallas Counties in Texas, and several places in Alaska. Additionally, there appear to be resurgences in several existing hot-spots so that their branches now become more distinguishable in the graph. Such places include Miami-Dade and Broward Counties in Florida and several Californian counties.

**Figure 9.**
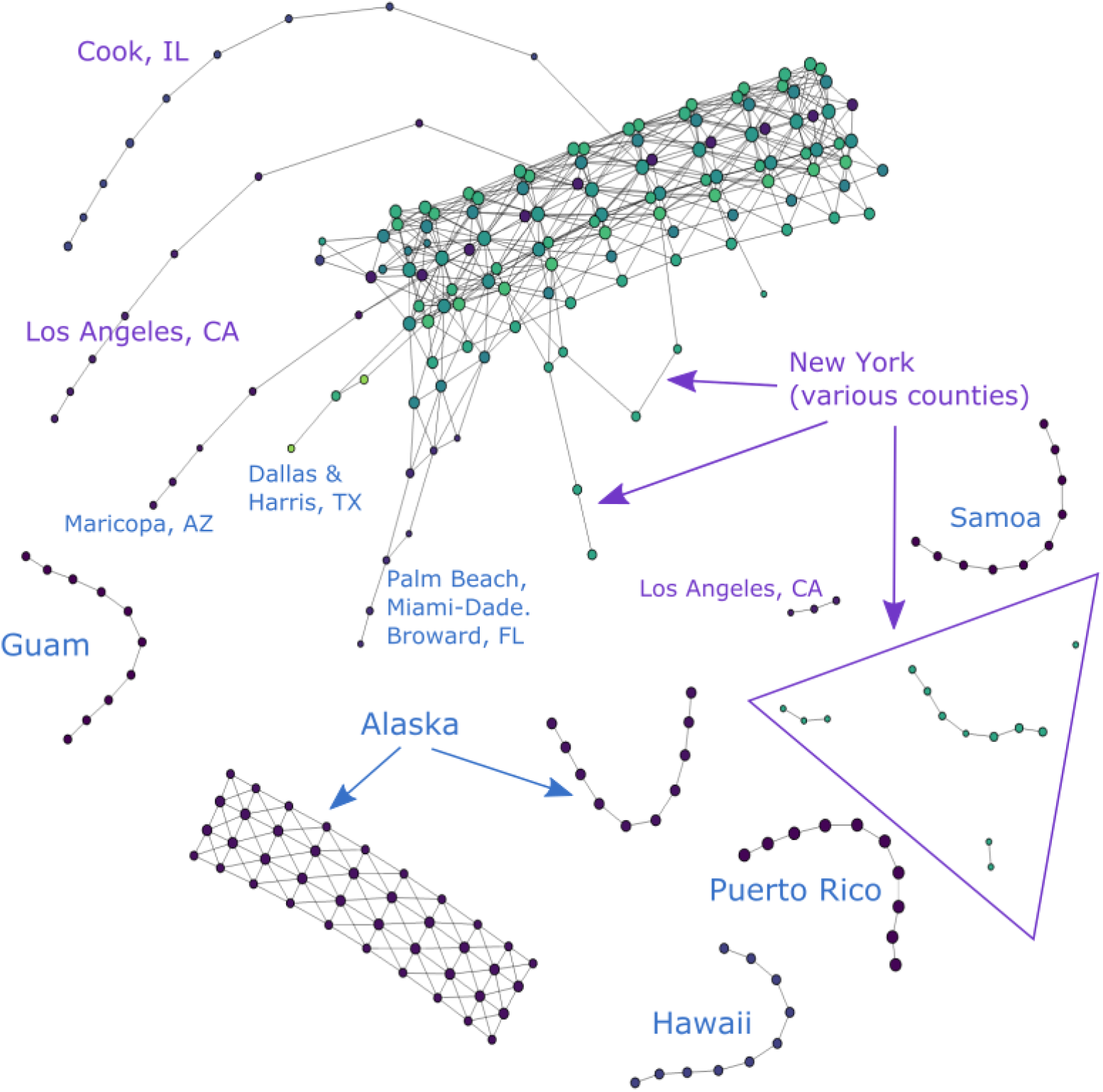
COVID-19 cases reported in the U.S. as of 7/24/20. (*n* = 10*, δ* = 8%)

**Figure 10.**
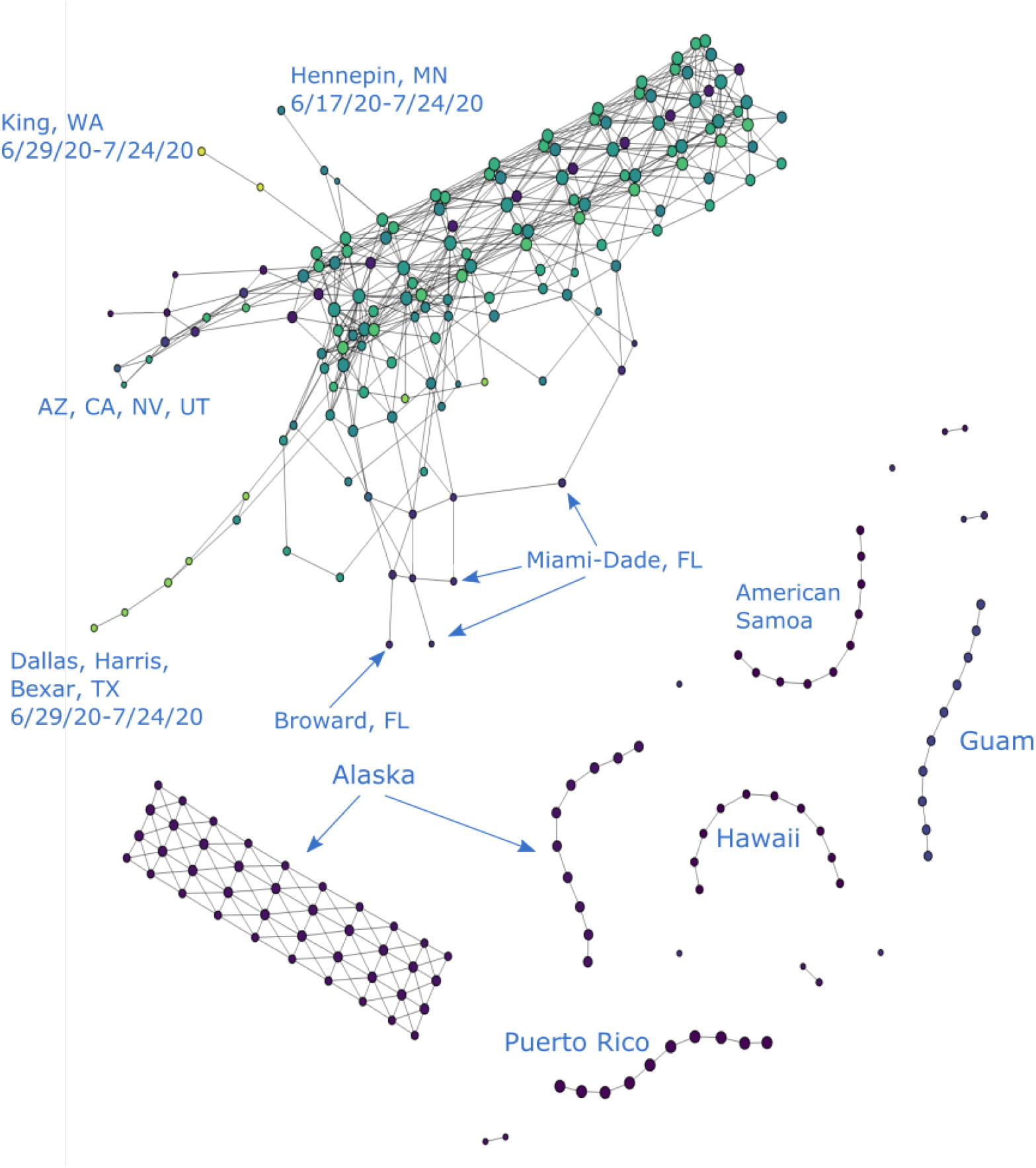
COVID-19 cases reported in the U.S. as of 7/24/20, excluding data from the entire New York State, Los Angeles, CA and Cook, IL. The four isolated nodes are Miami-Dade, FL for 7/8-7/11, 7/12-7/15, 7/16-7/18, and 7/19-7/22. Three of the four components consisting of two nodes and one edge are Maricopa, AZ for 7/1-7/5, 7/9-7/13, and 7/18-7/24. The fourth such component is Broward and Miami-Dade, FL for 7/5-7/21. (*n* = 10*, δ* = 8%)

From these updates, what we see is thus that, while embodying a coherent developmental progress of the pandemic, Mapper graphs are also in themselves fluid objects capable of change. We can get a more holistic picture of the entire progress by generating Mapper graphs with real-time data and follow its path of evolution. In the next section, we will discuss the evolution of Mapper graphs and the branches.

### 3.3. Evolution of Mapper graphs

Because our data incorporates temporal and geographic information, the resulting Mapper graphs are capable of conveying useful information regarding the gradual development of the COVID-19 pandemic across time and space. In Section 3.2, we studied this by inspecting different clusters as well as the branches sticking out of the main trunk of a particular Mapper graph. In this section, we demonstrate how one can obtain a more holistic picture of the growth of COVID-19 cases in the U.S. by studying the evolution of these features across Mapper graphs generated at different points in time.

For example, Figure 11 shows the evolution of Mapper graphs for unfiltered data through series of graphs generated from case data reported by 3/6/20, 4/10/20, 5/22/20, and 7/3/20, respectively. Figure 12 shows the evolution of Mapper graphs for filtered data. Since the number of total data points in each graph varies significantly, we adopt different resolution levels *n* and degrees of overlap *δ* to preserve relative visual coherence among graphs.

**Figure 11.**
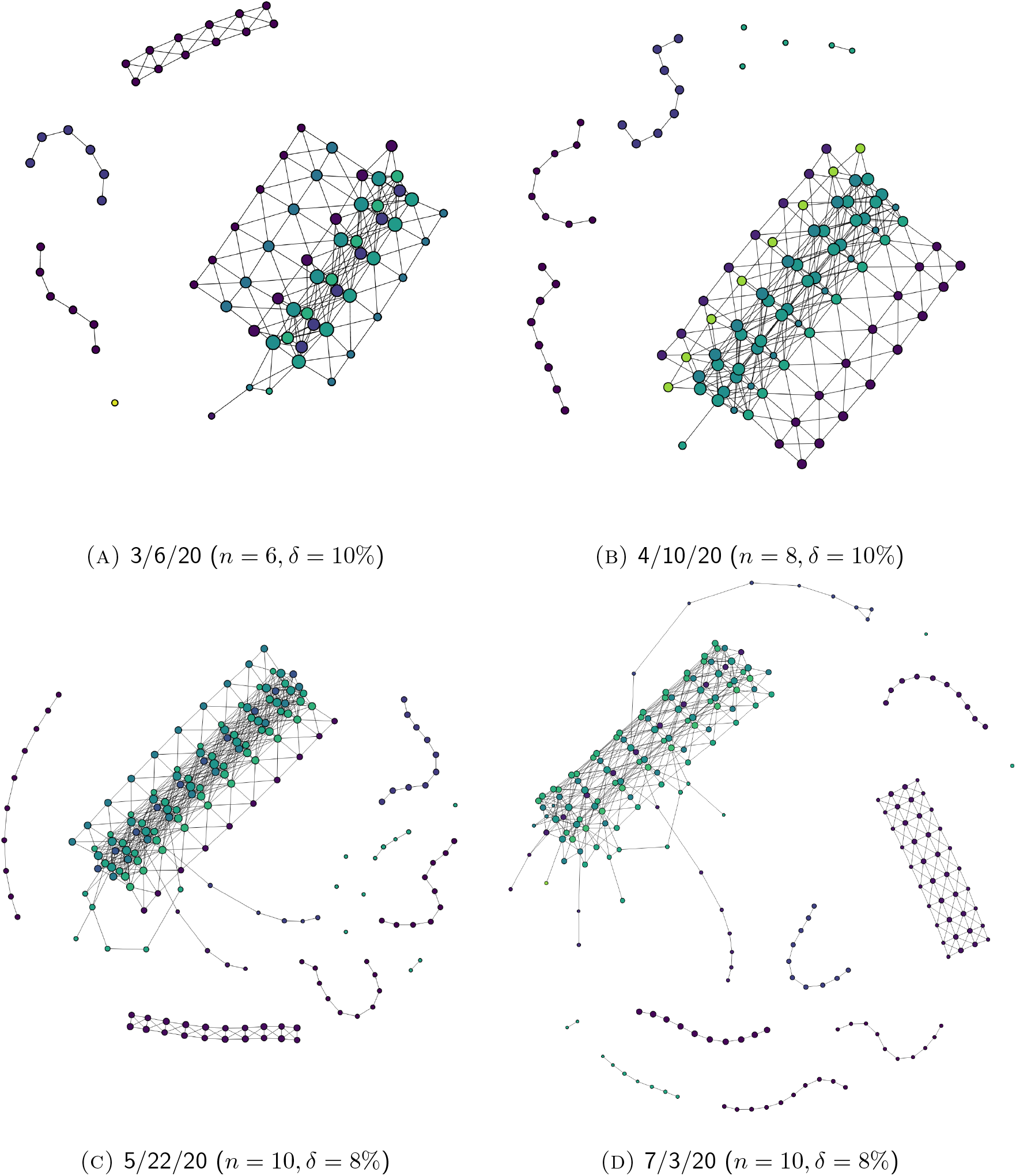
The evolution of Mapper graphs over time for unfiltered data.

**Figure 12.**
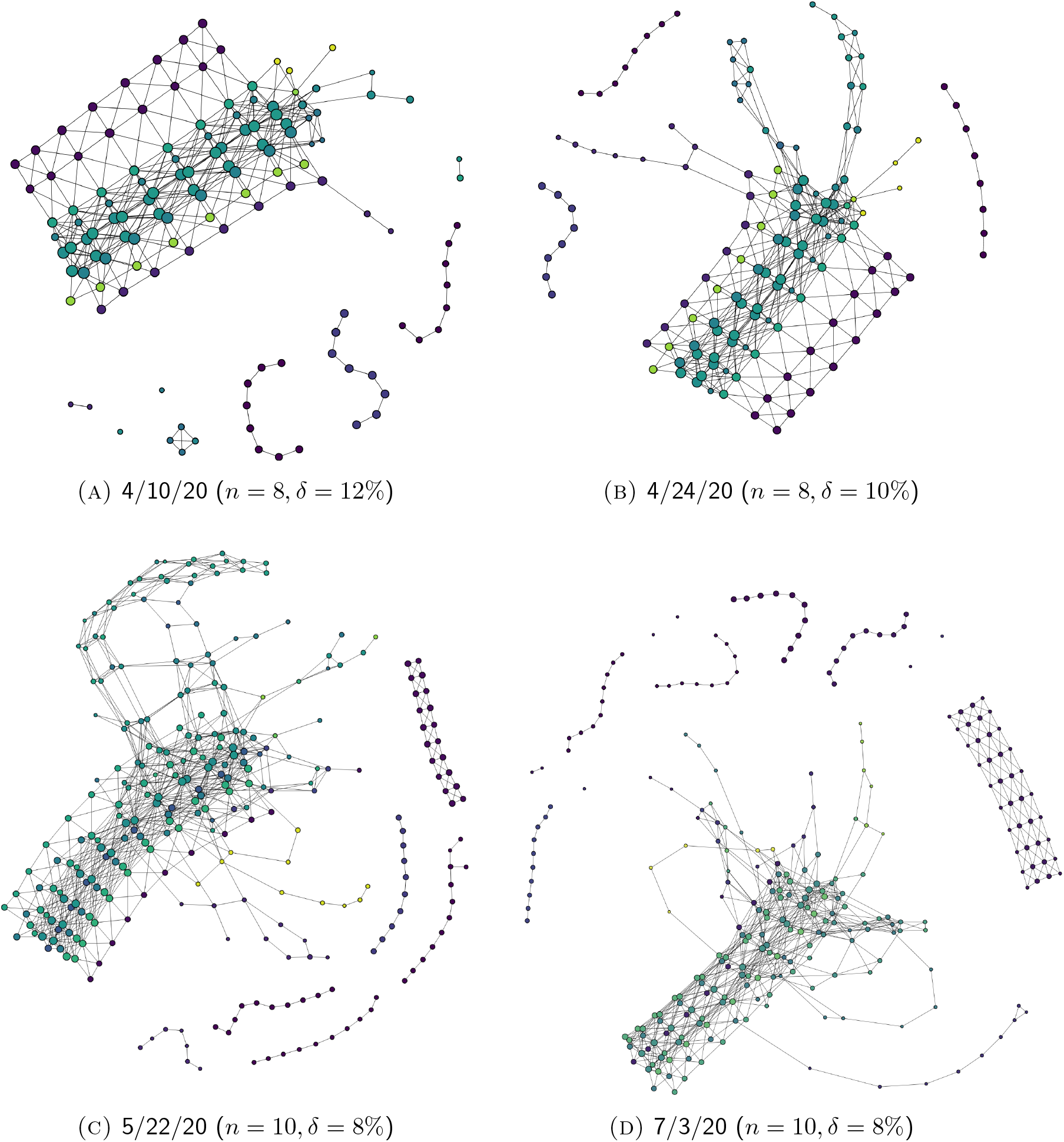
The evolution of Mapper graphs over time for filtered data (see Section 2.3.1).

As the number and distribution of U.S. COVID-19 cases evolve over time, the corresponding Mapper graphs evolve accordingly. New components might break off from the main trunk as the regions they represent experience aggressive increase in the number of reported cases. Existing isolated components might also reconnect to the main trunk through the creation of new edges due to a reduced disparity between the number of cases reported in the former and its neighboring regions. Additionally, branches might grow longer as new cases add up.

To illustrate these features, we will in the following sections shift the focus to a more local level and provide some detail on the evolution of Mapper graphs representing the local development of the pandemic in two places.

#### 3.3.1. New York State

Figure 13 gives a few evolutionary snapshots of the branch representing hot-spots in New York State. In earlier snapshots (Figures 13(A)–13(D)), the branch is unstable, with new disconnected nodes emerging frequently through each update so that the branch appears more fragmented. This reflects the acceleration of the pandemic in New York during that period. Each update in the data set brings significantly higher case numbers so that variability of projected data onto the “number of cases” dimension causes the break in the branch.

**Figure 13.**
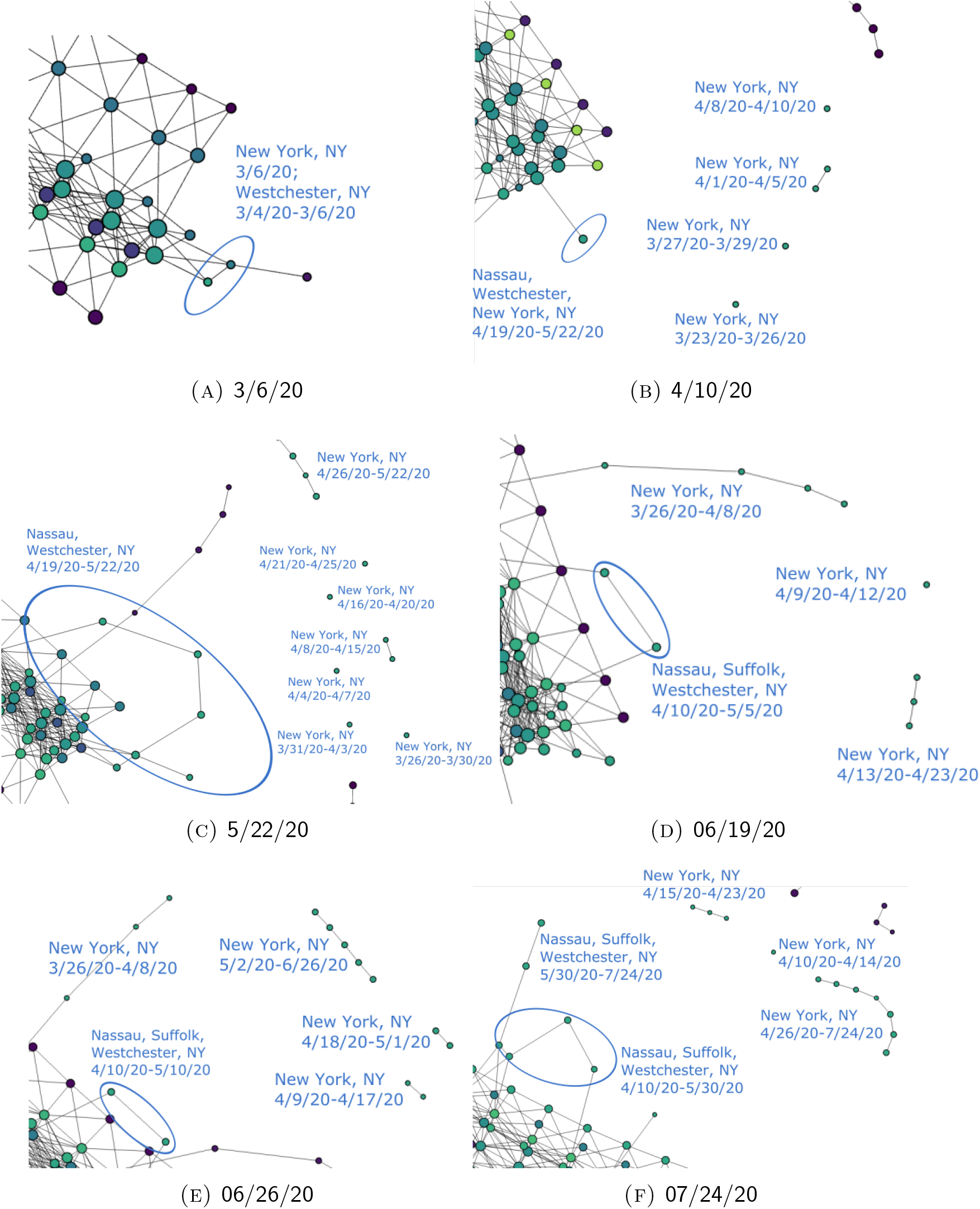
The evolution of the New York State cases over time.

In ensuing phases, however, the number of cumulative cases is so large that the relative relevance of each date’s numbers gets reduced as a result of the normalization (see Section 2.2.1). It thus becomes harder for new data to affect the shape of the branch. As a result, only the most impactful developments are picked up and reflected though every update. Hence, when the cases stabilize in New York, as we see from Figures 13(D)–13(F), the shape of its branch also tends to be less volatile. Instead of causing the branch to further break into more segments as in Figures 13(B) and 13(C), new nodes continue to build up on the latest segment. This shows that the pandemic no longer accelerates, notwithstanding a slowed growth. In the end, the structures that remain in the most recent graph, Figure 13(F), mark the most numerically relevant turning points in the development of the pandemic in New York, thus presenting a stable picture documenting the entire process.

In addition, it is also clear that, in contrast to earlier graphs in which New York is the only prominent branch, we can spot other branching structures in later graphs, easily discernible in Figure 11. This displays not only the development of the pandemic in other areas in parallel with New York, but also the declining relevance of New York on the national scale. Nevertheless, unlike other hot-spots whose branch almost disappeared from the graph because of their minimized relevance (e.g. most Connecticut hot-spots and King County in Washington), New York’s branch remains highly visible throughout the updates. This is due to the absolute numerical relevance of New York State’s data, which, by 7/23/20, ranks top in the U.S. in terms of state-level cumulative COVID-19 cases. Similarly, the branch representing California and Florida hot-spots also remain visible throughout the updates because of their absolute national relevance.

#### 3.3.2. Massachusetts-New Jersey complex

As elaborated in Section 3.2.2, because of their geographical proximity and national prominence, hot-spots in states like Massachusetts, Connecticut and New Jersey get reflected in the graph in the form of an entangled system of branches. The evolution of this system can be found in Figure 14. Similar to New York, these hot-spots have also gone through a process of acceleration and decline. In contrast, however, the size of this system is significantly smaller in later snapshots (see Figures 14(E) and 14(F)). This is because of the reduced national relevance of most of the hot-spots in this complex, so that their nodes no longer stand out of the main trunk as they are no longer distinguishable from their neighboring areas. As a result, only the most severe and most long-lasting hot-spots remained in this agglomeration, found among more recently developed systems of branches (see Figure 14(F)).

**Figure 14.**
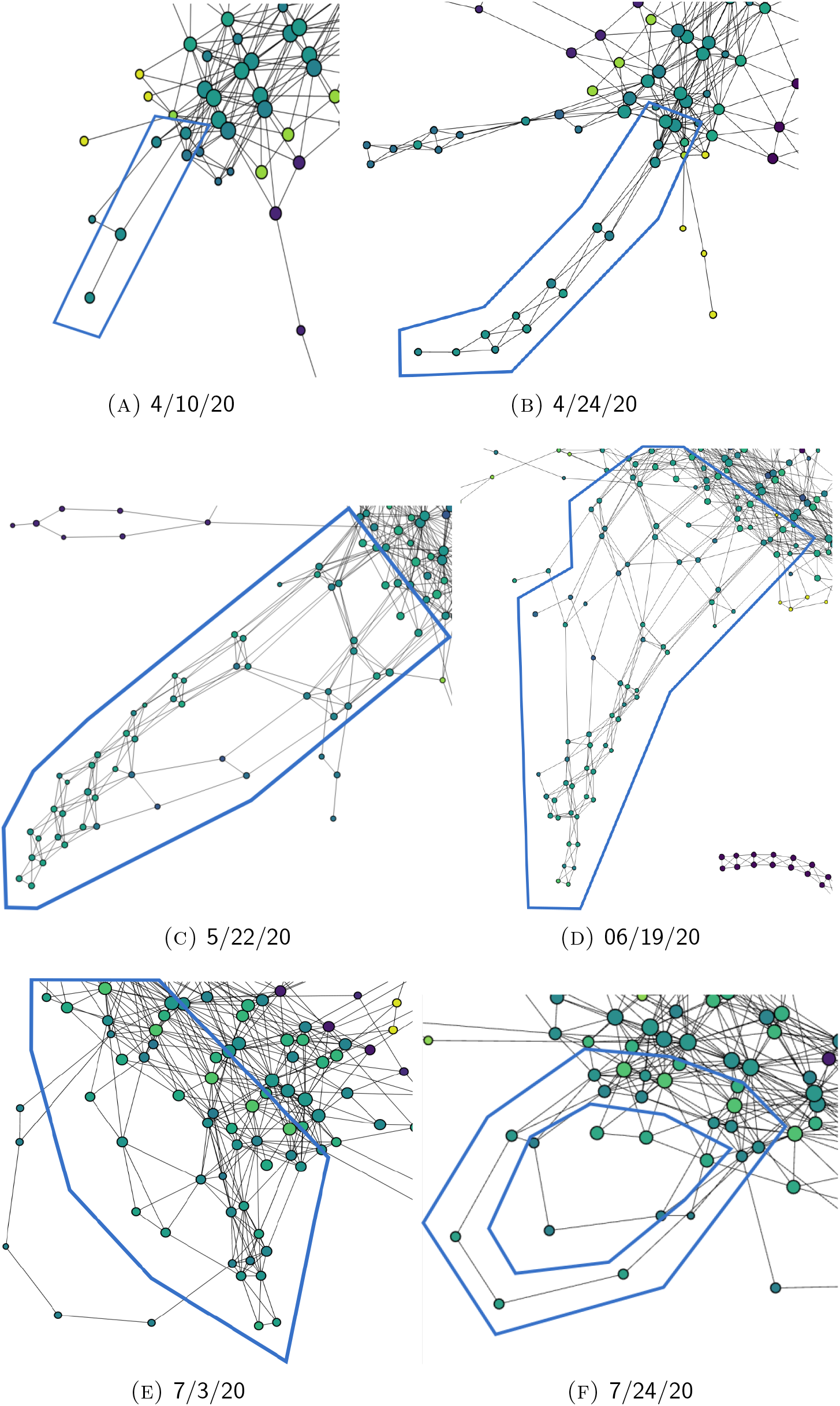
The evolution of the Massachusetts-New Jersey complex over time.

## 4. Conclusions and future work

The present paper provides a case study on the application of the Mapper algorithm to COVID-19 data collected in the United States. We have shown that Mapper captures a number of trends in the spread of COVID-19 and provides a more complete picture than those offered by more standard techniques of data visualization. The existence of (segmented) branches indicates hot-spots and the emergence of such branches correlates with troubling trends in the development of the pandemic in the regions they represent. Geographic proximity between hot-spots is also taken into account, so that branch complexes arise if nearby counties are similarly experiencing significant increase in the number of cases. The Mapper algorithm is also capable of tracking the gradual development of COVID-19 because it incorporates time data, supplying a more geographically and temporally complete picture of the spread of the virus. Continued observation on the evolution of the Mapper graphs is therefore clearly desirable.

There are various directions in which future analysis could be employed. For example, one could replicate the same study for data from different parts of the world. In addition, one can take into account border closures between countries, for example within EU or between EU and the rest of Europe.

One could additionally correlate stay-at-home orders issued by each U.S. state with critical changes observed in the Mapper graphs in hopes of evaluating the impact and effectiveness of such orders. Data about travel could also be included in hope that the Mapper reveals something new about how COVID-19 is spread by travelers through the U.S. In addition, one could overlay socio-economic data onto the Mapper graphs, along the lines of what was done in [5], providing important insight into how economic factors correlate to the spread of the disease across the U.S.

Unfortunately, the visualizations that we so far produced through the Mapper algorithm cannot offer strict and accurate predictions for the future development of the pandemic, since such predictions require the consideration of more real-life factors than we studied in this paper. But those factors can be incorporated by imposing more structure onto the Mapper. For example, we could define branches more rigorously using graph-theoretic notions. The length of the branches and the degree of the nodes in them can be assigned real-life meaning, along the lines of what was done by Escolar et. al. in [7]. This would provide more insight and understanding into the development of hot-spots, and could lead to a more predictive model given by the Mapper.

Another direction is to apply a different popular version of topological data analysis called *persistent homology*. In this setup, the data is turned into a topological space and one then studies the space’s connected components as well as “holes” of various dimensions. Persistent homology has been used to great effect in a number of settings, including biology, neuroscience, medicine, materials science, sensor networks, financial networks, and many others. For introductions to persistent homology and more details about its applications, see [1, 2, 6, 11]. The usefulness of persistent homology as a predictive epidemiological tool was demonstrated by Lo and Park [8] in their study of the Zika virus; emulating this approach might also lead to the construction of a useful predictive model for the coronavirus.

## Data Availability

The data is publicly available online.

https://systems.jhu.edu/research/public-health/ncov/

## Footnotes

1 Mapper was even used to analyze NBA player characteristics; see for example this article.

2 If one were to create the more complicated simplicial complex that carries even more information than the Mapper graph, then one would also look at all *n*-fold intersections for *n* > 1 to determine the *n*-simplices of the complex.

3 There are unclassified data labeled as “Out of state” or “Unassigned” in the original data set provided by CSSE. In order to preserve the coherency and usefulness of our analysis, we do not incorporate such data into our point cloud.

4 See here for more on DBSCAN.

5 For example, for a point cloud with data up to 6/21/20, the standard deviations for the four coordinates are calculated to be 0.0002336, 0.00022721, 0.0014227 and 0.00072409, respectively, after normalization.

6 Branches like these are sometimes called “flares” in the literature [7]. Our choice of terminology comes from the fact that we also have an evident “trunk.” In addition, “flaring” already has a meaning in the context of a spread of a virus.

7 This structure is unlikely to emerge in Mapper graphs for unfiltered data because New York data obscures the other minor hot-spots and makes them indistinguishable (see Section 2.2.1).

## Notes

### Competing Interest Statement

The authors have declared no competing interest.

### Funding Statement

No external funding for this research was received.

### Author Declarations

There is no need for approval for the research provided here since it only uses data that is publicly available.

